# Early sports specialization in Japanese young soccer players and related factors

**DOI:** 10.1101/2024.04.03.24305292

**Authors:** Yasuharu Nagano, Shogo Sasaki, Ayako Higashihara, Takashi Oyama

## Abstract

Although understanding the status of sports participation is essential for preventing injuries in young athletes, the level of specialization and relevant information in Japan is unknown. This study aimed to clarify the status of sports specialization and examine the relationships between specialization and training status in Japanese young soccer players.Four hundred Japanese young male soccer players were included. The players’ parents completed a web questionnaire that consisted of three-point questions regarding specialization and training status (year, days of playing soccer, age when starting soccer). The level of specialization and accompanying information were calculated, and data were compared by specialization status. Of the participants, 53.8% demonstrated a high level of specialization. In addition, 74.5% considered soccer more important than other sports, 89.0% trained in soccer for more than 8 months of the year, and 74.0% had quit other sports to focus on soccer or played only soccer. The proportion of participants who played only soccer was significantly higher in the high-specialization group (37.6%) than in the moderate-specialization (22.5%; *P* < .01) and low-specialization (7.1%; *P* < .01) groups. By specialization status at grades 4 to 6 (9–12 years), 40.3% of participants demonstrated a high level of specialization. Young Japanese soccer players tend toward early specialization. Factors contributing to the high-specialization level are being active throughout the year and rarely playing other sports. Training volume should be controlled in children of this age with avoidance of early specialization.

## Introduction

For children, soccer is efficacious in improving physical capacity, health-related fitness parameters, and self-esteem.[1] Soccer has been a highly popular sport for young athletes. Meanwhile, early specialization in a particular sport and increased participation in organized sports with increased training volume have become prevalent for improving athletic performance and achieving social success. However, a previous meta-analysis found that sport specialization in athletes younger than 18 years was associated with an increased risk of musculoskeletal overuse injuries.[2] Therefore, position statements from sports medicine organizations have generally been consistent in recommending against early sport specialization.[3] To provide fundamental information for injury prevention in young athletes, specialization status must be clarified.

Sports specialization is defined as “intense, year-round training in a single sport at the exclusion of other sports.”[3, 4] To evaluate the degree of sports specialization, the three-point Jayanthi scale [3, 4] is popularly used and is based on three key components: year-round training, choosing a main sport, and quitting other sports. Previous studies have reported on the specialization status of young athletes. Post et al [5] reported that among all youth athletes (mean age 14.2 years), 41.8% had high-specialization status, and this proportion was 35.3% among soccer players. Post et al [6] also reported that the status of high sports specialization by age in young athletes including soccer as follows: 30.2% at 12 years of age, 38.1% at 13 years of age, and 42.3% at 14 years of age. Bell et al [7] reported that the proportion of high-specialization status of youth male athletes including soccer (mean age 15.7 years) was 36.9%. However, there have been no reports of the sports specialization status of Japanese young athletes.

One reason for specialization is the status of other sports played. Although a previous study showed a high percentage of young students (77.9%) played multiple sports,[7] in Japan, the proportion of children who play multiple sports is very low, ranging from 10.0% [8] to 17.4% [9] for children in grades 7–9 and 22.9% for soccer players.[9] In addition, some athletes have only ever played a single sport, and athletes who continued a single specific sport from elementary school age are more likely to have overuse disorders, especially in team sports.[10] Consequently, it is possible that young Japanese athletes are highly specialized due to a low percentage of multiple sports and that their specialization is increasing as they have no or little athletic experience other than soccer.

Another factor related to the level of specialization is training volume. In young soccer players, training volume of >5 days per week playing soccer was related to overuse injuries.[5] In some recent studies in Japan, the number of days of weekly activity was related to acute injuries and overuse injuries during grade 7–9.[9] In addition, the number of training days per week was related to growth-related knee and heel pain in Japanese junior soccer players.[11] Other studies have focused on the number of months of sports per year, with more months spent playing sports being associated with acute injuries and overuse injuries.[5] Although it is assumed that training volume increases with the level of specialization, the relationship between training volume and the level of specialization in current young Japanese soccer players remains unclear.

Although understanding the status of sports participation is essential to prevent injuries in young athletes, the level of specialization and relevant information in Japan remains unknown. The purpose of this study was to clarify the status of sports specialization and examine the relationships between specialization and training status (only sports carrier of soccer, days spent playing soccer, and age when starting soccer) among Japanese young soccer players. Our first hypothesis is that the specialization status of Japanese young soccer players is higher than that reported in previous studies and related to only sports carrier of soccer. Our second hypothesis is that high sports specialization of Japanese young soccer players is related to more days per week of playing soccer.

## Materials and Methods

### Study setting

This study was a cross-sectional study of data obtained from a web-based questionnaire. The questionnaire was randomly distributed to people registered with a web questionnaire supplier (Rakuten Insight Inc.). The aim was to recruit 400 subjects whose male child played soccer and attended school in grades 8 (13–14 years) or 9 (14–15 years). According to the power analysis,[12] the required sample size for a descriptive study of a dichotomous variable (*P* = .40, W = 0.10, confidence level = 95%) was 369. Based on this value, we targeted a sample size of 400. We recruited an equal number of participants from grades 8 and 9. First, we randomly distributed a screening questionnaire to 13,852 people registered with a web questionnaire supplier. A total of 6,099 of the 13,852 individuals participated in the screening survey. Next, based on the results of the screening survey, 723 parents of young male soccer players in the seventh to ninth grades participated in the main survey. Finally, 6 data points were excluded through data cleaning, and 600 out of the 717 collected datasets, with 200 from each school grade, were randomly selected by a questionnaire supplier. A series of surveys achieved the desired sample size in 3 days (August 1–3, 2022). Although parents of seventh-grade players also responded to the survey, they were excluded from this study because their schools had changed from elementary to junior high in the previous year, and their soccer-playing environment had largely changed at the half-year. All respondents provided informed consent by displaying and clicking on a screen before participating in the study. Respondents who agreed to participate in this survey answered the questionnaire voluntarily, and information was collected without revealing the identity of any individual participant to the researcher. The ethical review board of the authors’ institution approved the present study. This research was conducted based on the principles of the Declaration of Helsinki.

### Electronic questionnaire

The questionnaire consisted of four screening questions and six main questions (Table 1). The screening survey assessed whether the respondent had a child with the following criteria: (1) currently playing soccer in the club team, (2) in seventh to ninth grades, (3) male, and (4) playing soccer regularly for more than 1 day per week. If answers to the screening questions matched the inclusion criteria, the questionnaire continued to the next main survey. If the participant did not match even one of the inclusion criteria, the respondents were excluded from the survey. The main questions asked respondents about their sports activity (year, age at which they started playing soccer, participation in sports during seventh to ninth grades and during fourth to sixth grades), and status of specialization during the seventh to ninth grades and fourth to sixth grades. Status of sport specialization was determined with a widely used 3-point specialization scale.[3, 4] This scale was calculated from a series of three questions that asked (1) if the athlete quit other sports to focus on soccer, (2) if they viewed soccer as more important than other sports, and (3) if they trained or participated in soccer more than 8 months of the year.[13] Specialization was classified as “high” for those who met all three questions, “moderate” for those who met two, and “low” for those who met only 0 to 1. Those who answered “No, plays only soccer” to question 1 (Q6) were considered to meet this criterion.

**Table 1:**
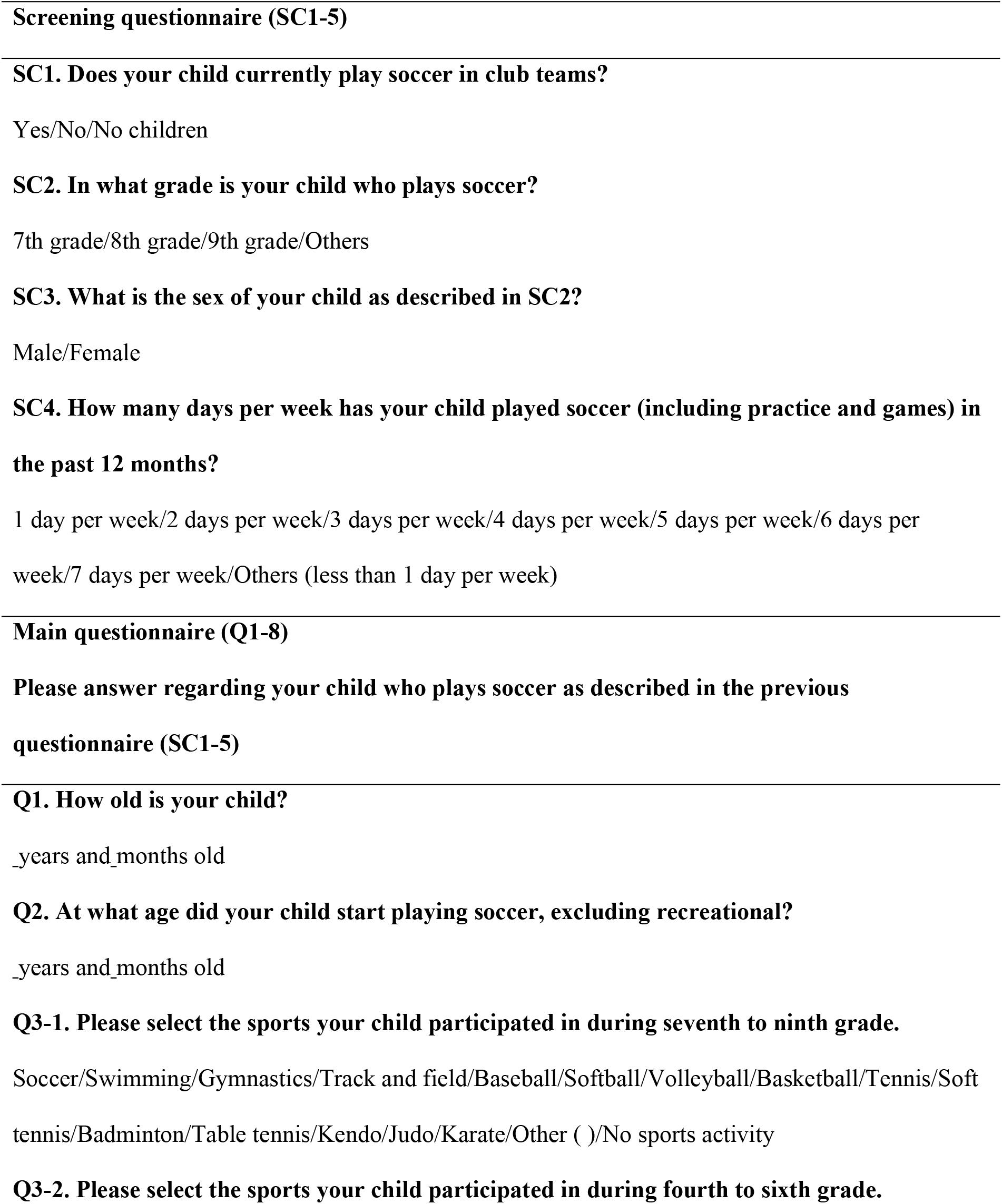
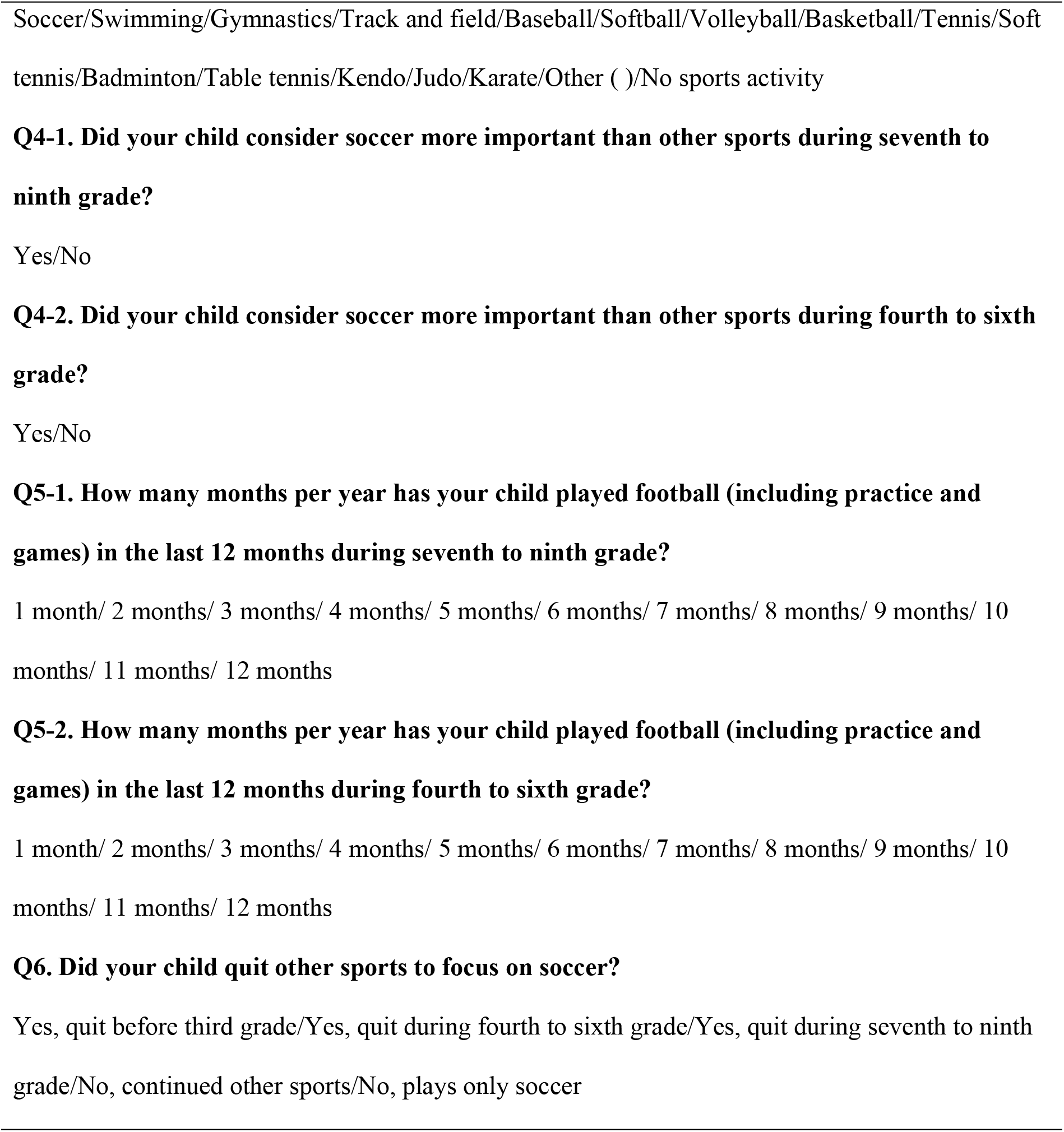
Questionnaire regarding sports specialization and related factors in young male soccer players.

### Statistical analysis

We summarized the data as means and standard deviations, frequencies and proportions (%) by specialization status. Using chi-square test, we compared the proportion of those who play only soccer by specialization status. Days of playing soccer per week and age starting soccer were compared by specialization status using one-way analysis of variance, along with Bonferroni correction performed as a post hoc test. All statistical analyses were performed using SPSS Statistics version 19.0 for Windows (IBM; Brush Prairie, WA, USA), and results were considered statistically significant with an α level of *P* < .05.

## Results

A total of 400 participants (mean age, 13.9 ± 0.9 years) completed the questionnaire. Table 2 presents the proportion of participants classified as high, moderate, and low specialization: 53.8% of participants demonstrated a high level of specialization, 32.3% demonstrated a moderate level of specialization, and 14.0% of participants demonstrated a low level of specialization. Among the participants, 74.5% considered soccer more important than other sports, 89.0% trained for more than 8 months of the year in soccer, and 74.0% had quit other sports to focus on soccer or playing only soccer. In total, 81.5% of participants and 92.1% of the high-specialization group trained 12 months of the year. The proportion of participants who playing only soccer was significantly higher in the high-specialization group (37.6%) than in the moderate-specialization (22.5%; *P* < .01) and low-specialization (7.1%; *P* < .01) groups, and it was significantly higher in the moderate-compared with the low-specialization group (*P* < .01). The number of days spent playing soccer per week was significantly greater in the high-specialization group (4.6 ± 1.2 days, mean ± SD) than in the moderate- (4.1 ± 1.5 days; *P* < .01) and low-specialization (3.8 ± 1.7 days; *P* < .01) groups. The age at which the participant started playing soccer was significantly higher in the high-specialization group (7.8 ± 2.7 years) than in the moderate-specialization group (9.0 ± 2.9 years; *P* < .01). In specialization status at grades 4 to 6 (9–12 years), 40.3% of participants demonstrated a high level of specialization, and 71.6% of the high-specialization group already demonstrated a high level of specialization.

**Table 2.** Frequency of sports participation and prevalence of injury.

|  | Specialization Status |  |  |  |  |  |  |  |
| --- | --- | --- | --- | --- | --- | --- | --- | --- |
|  | Low |  | Moderate |  | High |  | Total |  |
| Number | 56 | (14.0%) | 129 | (32.3%) | 215 | (53.8%) | 400 | (100.0%) |
| Soccer more important |  |  |  |  |  |  |  |  |
| Yes | 3 | (5.4%) | 80 | (62.0%) | 215 | (100.0%) | 298 | (74.5%) |
| No | 53 | (94.6%) | 49 | (38.0%) | 0 | (0.0%) | 102 | (25.5%) |
| Months of participation |  |  |  |  |  |  |  |  |
| 1 | 8 | (14.3%) | 1 | (0.8%) | 0 | (0.0%) | 9 | (2.3%) |
| 2 | 0 | (0.0%) | 2 | (1.6%) | 0 | (0.0%) | 2 | (0.5%) |
| 3 | 3 | (5.4%) | 4 | (3.1%) | 0 | (0.0%) | 7 | (1.8%) |
| 4 | 4 | (7.1%) | 2 | (1.6%) | 0 | (0.0%) | 6 | (1.5%) |
| 5 | 1 | (1.8%) | 3 | (2.3%) | 0 | (0.0%) | 4 | (1.0%) |
| 6 | 7 | (12.5%) | 6 | (4.7%) | 0 | (0.0%) | 13 | (3.3%) |
| 7 | 1 | (1.8%) | 2 | (1.6%) | 0 | (0.0%) | 3 | (0.8%) |
| 8 | 1 | (1.8%) | 2 | (1.6%) | 4 | (1.9%) | 7 | (1.8%) |
| 9 | 1 | (1.8%) | 1 | (0.8%) | 3 | (1.4%) | 5 | (1.3%) |
| 10 | 1 | (1.8%) | 5 | (3.9%) | 6 | (2.8%) | 12 | (3.0%) |
| 11 | 0 | (0.0%) | 2 | (1.6%) | 4 | (1.9%) | 6 | (1.5%) |
| 12 | 29 | (51.8%) | 99 | (76.7%) | 198 | (92.1%) | 326 | (81.5%) |
| Other sports |  |  |  |  |  |  |  |  |
| Playing only soccer | 4 | (7.1%) | 29 | (22.5%)* | 83 | (38.6)** | 116 | (29.0%) |
| Quit before grade 3 (–9 y) | 2 | (3.6%) | 8 | (6.2%) | 36 | (16.7%) | 46 | (11.5%) |
| Quit at grades 4–6 (9–12 y) | 1 | (1.8%) | 24 | (18.6%) | 73 | (34.0%) | 98 | (24.5%) |
| Quit after grade 7 (12 y–) | 5 | (8.9%) | 8 | (6.2%) | 23 | (10.7%) | 36 | (9.0%) |
| Continuing other sports | 44 | (78.6%) | 60 | (46.5%) | 0 | (0.0%) | 104 | (26.0%) |
| Days of playing soccer,<br>mean (SD), d/wk | **3.8** | **(1.7)** | **4.1** | **(1.5)** | **4.6** | **(1.2)\*\*** | 4.3 | (1.4) |
| Age starting soccer, mean<br>(SD), y | 8.8 | (3.0) | 9.0 | (2.9) | **7.8** | **(2.7)#** | 8.3 | (2.8) |
| Specialization status at<br>grades 4–6 (9–12 y) |  |  |  |  |  |  |  |  |
| Other sports | 16 | (28.6%) | 25 | (19.4%) | 21 | (9.8%) | 62 | (15.5%) |
| Low | 36 | (64.3%) | 15 | (11.6%) | 7 | (3.3%) | 58 | (14.5%) |
| Moderate | 4 | (7.1%) | 82 | (63.6%) | 33 | (15.3%) | 119 | (29.8%) |
| High | 0 | (0.0%) | 7 | (5.4%) | 154 | (71.6%) | 161 | (40.3%) |

Values in bold font indicate significantly different between groups.
\*Significantly different from low level of specialization ( $P < .01$ ).
\*\*Significantly different from low and moderate level of specialization ( $P < .01$ ).

## Discussion

The results of this study showed that 53.8% of young Japanese soccer players had a high-specialization level, and 40.3% were already at a high level of specialization when they were in grades 4 to 6. Factors that contributed to the high-specialization level were being active throughout the year and rarely continuing to play other sports. We found that, also related to the high-specialization level, the athletes had no previous experience in sports other than soccer, and they were active more days per week. These results were consistent with our hypothesis. The level of specialization of young Japanese athletes has not been previously determined, and the results of this study may provide a baseline for comparing the sporting environment of young athletes with the findings of other studies.

The level of specialization of young Japanese soccer players in this study was higher than in previous research.[5-7] In addition, their level of specialization in grades 4–6 was similar to that reported in previous studies with older participants.[5, 6] Because high specialization is associated with the occurrence of overuse injuries,[14] the high specialization of young Japanese soccer players potentially threatens their health status. In fact, the incidence of growth-related injuries among young Japanese soccer players [11, 15] is higher than that reported in a previous study from another country.[16] Osgood–Schlatter disease, which is most likely to be a problem in soccer players, tends to occur at the age of this study’s participants.[17] Soccer-related motions, including kicking, running, landing, and cutting, place a substantial load on knee extension, and the repetition of these loads is associated with a higher risk of developing Osgood–Schlatter disease.[18] To prevent these injuries, excessive training associated with early specialization must be avoided.

One reason for the high level of specialization was the number of months of activity per year. Although the criterion for specialization is ≥8 months of activity per year,[3, 4] most of the participants in present study were active throughout the year. Sports activities for young players in Japan are typically conducted year-round rather than seasonally, and the present study clearly illustrates this situation. The number of months active per year was also related to the occurrence of overuse disorder,[6] but it is not realistic to control the number of months active in a situation in which most players are active throughout the year. The results of the present study showed significant differences in the number of weekly activity days between the levels of specialization, which might be useful for controlling the amount of training.

Another reason for the high level of specialization was the low implementation of playing sports other than soccer. Only 26% of the participants continued to play other sports, which is much lower than the proportion reported in the previous study (77.9%).[7] Furthermore, approximately 30% of the participants had no experience in sports other than soccer, and 38.6% of those with a high level of specialization found themselves in a situation where they had no experience in sports other than soccer. Meanwhile, although the age at which participants started playing soccer was similar to that in a previous study,[6] those with a high level of specialization started soccer at an earlier age. This means that in Japan, especially among the U12 and U15 age groups, soccer is immensely popular, with numerous teams available,[19] resulting in a strong tendency for specialization to occur at an earlier stage in a player’s sports journey. From the perspective of long-term athlete development,[20] children are encouraged to engage in multiple sports, especially team ball sports, as early specialization in a single sport can result in an increased risk of overuse injuries in later youth stages.[3, 10] However, athletes who play multiple sports are at risk of increasing their total training volume,[9] which should be controlled.

This study has several limitations. First, this was a cross-sectional study with a self-reported questionnaire in which parents recalled previous information. Thus, there is the possibility of recall bias in the results. However, we assumed that information on sports experience over the past several years can be collected satisfactorily. Second, the relationships between sports specialization and success or performance in the sports were not evaluated. Finally, we did not examine the relationship between sports specialization and the occurrence of injuries. The influence of sport specialization on injury occurrence and future injury risk in soccer players should be examined in the future. However, the result of the present study can provide a foundation for these future studies.

## Conclusion

We clarified the status of sports specialization and the relationships between specialization and training status in Japanese young soccer players. Japanese young soccer players have a tendency toward early sports specialization. Factors that contributed to the high-specialization level were being active throughout the year and rarely continuing to play other sports. The training volume should be controlled in children of this age, with early specialization should be avoided.

## Data Availability

All relevant data are uploaded to Zenodo at https://doi.org/110.5281/zenodo.10874130 from the time of publication.

